# Evaluating the influence of sleep quality and quantity on glycemic control in adults with Type 1 Diabetes

**DOI:** 10.1101/2022.07.20.22277229

**Authors:** Marta Botella-Serrano, J. Manuel Velasco, Almudena Sánchez-Sánchez, Oscar Garnica, J. Ignacio Hidalgo

## Abstract

**Background:** Sleep quality disturbances are frequent in adults with type 1 diabetes. However, the possible influence of sleep problems on glycemic variability has not been deeply studied in the past. This study aims to assess the impact of sleep quality and sleep quantity on glycemic control.

**Materials and Methods:** Observational study in 25 adults with type 1 diabetes, simultaneous recording of continuous glucose monitoring (Abbott FreeStyle Libre system) for 14 days, and a sleep study by wrist actigraphy (Fitbit Ionic device). The study analyzes, using artificial intelligence techniques, the relationship between the quality and structure of sleep with time in normo-, hypo-, and hyperglycemia ranges and with glycemic variability. The patients are also studied as a group, comparing patients with good and poor sleep quality. Several cluster analyses and correlational studies are performed

**Results:** A total of 243 days/nights were analyzed, of which 77% (n=189) were categorized as poor quality and 33% (n=54) as good quality. Linear regression methods find a correlation (r=0.8) between the variability of sleep efficiency and the variability of mean blood glucose. With clustering techniques, patients were grouped according to their sleep structure (characterizing this structure from the number of transitions between the different sleep phases). These clusters show a relationship between time in range and sleep structure.

**Conclusions:** This study suggests that poor sleep quality is associated with lower time in range and greater glycemic variability, so improving sleep quality in patients with type 1 diabetes could improve their glycemic control.

## 1. Introduction

Alterations in the quantity and quality of sleep are common in the general population and people with Type 1 Diabetes Mellitus (T1DM).^1, 2^ In T1DM people, a shorter duration of the deep sleep phase has been demonstrated in both adults and children,^3, 4^ and the poor subjective quality of sleep, excessive daytime sleepiness,^1, 2^ and higher prevalence of obstructive sleep apnea.^5^

The impact of these disturbances on glycemic control in adults with type 1 diabetes has been little studied. However, previous studies suggest that sleep disturbances decrease insulin sensitivity, worsen glycemic control, and increase glycemic variability.^6^

The main objective of this study is to analyze in a group of patients with type 1 diabetes the impact of sleep disturbances on short-term glycemic control, glycemic variability, and the frequency of hypoglycemia. For this purpose, flash continuous glucose monitoring (performed by Abbott FreeStyle Libre devices) and a sleep study using wrist actigraphy (Fitbit Ionic device) were carried out simultaneously for 14 days in a set of patients. The CGM data includes interstitial blood glucose levels recorded during the entire time the patient wore the sensor, not only during sleep. Twenty-five patients were included, of which data from three patients had to be discarded, with a total of 243 nights/days recorded. The study analyzes interindividual differences in glycemic control, in relation to nights with worse or better quality/quantity of sleep.

Nocturnal patterns have also been studied as a group, comparing nights with good and poor sleep quality.

## 2. Materials and Methods

### 2.1. Study Design and Patient Selection

#### 2.1.1 Inclusion Criteria

- Patients aged between 18 and 65 years.
- T1DM with at least one year of duration.
- In treatment with an insulin pump or multiple doses of subcutaneous insulin per day (MDI).
- Availability of a mobile phone with direct scanning capability for reading the FreeStyle Libre system sensor.
- They understand and give informed consent to participate in the study.

#### 2.1.2 Exclusion criteria

- Diagnosis of major psychiatric disorder.
- Treatment with corticoids.
- Ketoacidosis that required hospital admission in the last six months.
- Severe intercurrent disease in the last six months.

#### 2.1.3 Procedures

- Pre-screening Visit: The study was explained to the participants. All patients signed a prior informed consent form. Patients continued with their usual treatment. Sociodemographic variables, anthropometric data, and clinical data were collected from the patients’ medical records.
- Visit 1: Patients were given a wristband with wristwatch actigraphy (Fitbit Ionic model). Free Style free sensor was placed for continuous glucose monitoring for 14 days. The sensor was connected to the Abbot Libre View platform. Patients self-completed two surveys:
  - Pittsburg survey to assess habitual self-perceived sleep quality (PSQI index).^1^
  - Munich survey to assess social jet lag.^7, 8^
- Visit 2: The glucose sensor and wrist actigraphy were removed.

### 2.2. Data gathering and preprocessing

Recording of blood glucose data was performed automatically through the Abbott Libre View application. Time in the range is estimated directly by the FreeStyle Libre systems. In addition, it is calculated from the microdata generated by the meter using the Rosendaal method,^9^ which assumes a linear progression between two glucose values and calculates the specific value for each minute (linear interpolation). The same process is used to calculate time in hypo- and hyperglycemia. The recording of the wrist activity is also performed automatically and digitized and can be visualized from a web page. Detailed information is recovered from the Fitbit site using own software developed in python.

### 2.3. Sleep monitoring variables to be analyzed

The sleep monitoring variables analyzed in this study are:

- Time in bed (Bed). Total time spent in bed, both awake (Awake) and asleep (Asleep).
- Duration of light sleep (Light).
- Duration of deep sleep (Deep).
- Duration of REM phase (REM).
- Sleep efficiency (Efficiency): the ratio of total sleep time to time in bed.^10^
- Average sleep duration/ day (average of last 15 days).
- The number of REM sleep episodes.
- Temporal distribution of sleep phases.
- Number and duration of micro awakenings.

### 2.4. Glucose monitoring variables to be analyzed

The glucose monitoring variables analyzed in this study are

- Time in range (70-180 mg/dl) (TR).
- Mean blood glucose (mg/dl) (Mean_glucose).
- Standard deviation (SD).
- Coefficient of variation (CV).
- Time in level 1 hypoglycemia (54-70 mg/dl) and level 2 hypoglycemia (*<* 54 mg/dl) (T Hyper).
- Time in hyperglycemia level 1 (180-250 mg/dl) and level 2 (*>* 250 mg/dl) (T Hypo).
- Hypoglycemia/hyperglycemia episodes (minimum duration 15 min).
- Mean Amplitude of Glycemic Excursions (MAGE).
- Mean Daily Glucose Differences (MDGD).

### 2.5. Statistical Methods

To find out the possible relationship between the glucose levels of the patients under study and the quality of sleep, several cluster analyses and correlational studies are performed using the R language and related libraries.^11, 12^

These types of models, together with language processing techniques taken from the field of artificial intelligence, make it possible to find possible patterns of glucose under the different variables selected and the nocturnal sleep patterns recorded for each of the patients.^13^

The K-means algorithm is used in the various cluster analyses.^14^ First, considering only the variables associated with sleep, then the variables associated with glucose levels, and, finally, the set of all variables. The correlation analysis (see section 3.3) takes as a reference Pearson’s correlation coefficient, which, with values close to 1 (−1), confirm the existence of a positive (negative) linear relationship between the corresponding variables.

#### 2.5.1 Sequence of Sleep States

Figure 1 shows the workflow of the methodology. On the one hand, we record the daily time series of blood glucose levels of a set of individuals using Free Style Libre sensors. At the same time, we register the sleep state sequences of these individuals during the corresponding nights. Throughout the night, the person transits between different sleep states (wake, light, rem, deep) forming a time series of states or categories.^12^ Figure 1 (left-top) presents a visual representation of this time series as a sequence of colors displaying the sleep states. In Figure 1 we can observe that patient 1 (HUPA001), one night went through 33 different states, following a sequence: wake, light, wake, light, light, deep, etc.

**Figure 1:**
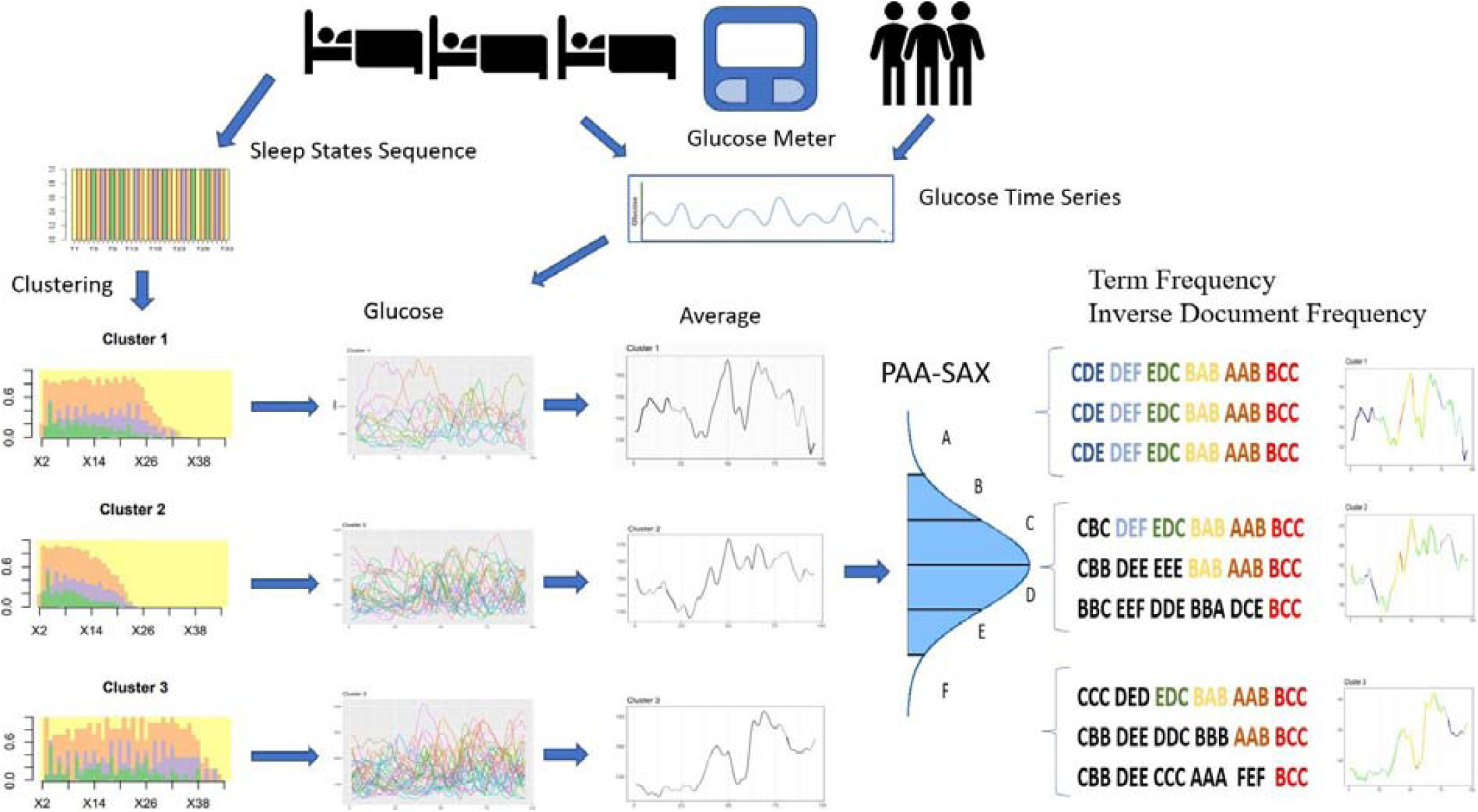
Design flow of the methodology presented in this paper.

In this study, we want to determine if we can distinguish different patterns of sleep time series to subsequently find if there is a correlation between these sleep patterns during the night and the patterns of blood glucose levels evolution throughout the day. To search for sleep patterns, we use different time series clustering techniques.^15^ Using all the data from all patients, we can group the sleep state sequences based on their similarity,^12^ i.e., each grouping (or cluster) includes those time sequences that are most similar. In Figure 1, we can see that a grouping example in three clusters, i.e., three patterns of the sleep behavior of the participants in this study. After performing the clustering, we group the daily time series of glucose levels corresponding to the nights of each cluster. These glucose time series are averaged, and we obtain the evolution of the glucose level characteristic of each cluster (i.e., of each sleep behavior pattern).

### 2.6. Similarity among Glucose Time Series

From the clusters obtained from the overnight sleep states, we can group the time series of the blood glucose level for the corresponding days and obtain the average of these (Figure 1 bottom-center-left). These figures give us an idea of the average glucose behavior for each sleep cluster. To identify specific behaviors of each cluster, we use techniques commonly used in language processing; notably those described in Zan et al.^16, 17^ To do this, we transform a time series of numerical glucose values into a sequence of characters that is similar to words.

First, we transform the numerical values of blood glucose into a sequence of symbols. To obtain these symbols, (Figure 1 bottom-center-right), the time series is normalized and reduced by obtaining the average of a number *n* of glucose values (in this work, *n* = 4) (this technique is called Piecewise Aggregate Approximation or PAA),^18^ and each average point is assigned a symbol within a dictionary based on the statistical distribution (Symbolic Aggregate approXimation or SAX).^16, 17^ Finally, the symbols are grouped into words of a specific size (12 in this work).

Next, we use a well-known technique in language processing that identifies the topics covered in a set of documents from the frequencies of occurrence of words in each document and in the set of documents. In this way, we seek to identify behaviors specific to each cluster and those common to all clusters. In this work, a cluster is equivalent to a “document.” In the bottom-right of Figure 1, we can see the process summarized and simplified using words of size 3. Based on the number of occurrences of each “word” in all the time series of each cluster, we obtain the weight vectors associated with each word (Term Frequency Inverse Document Frequency^19^). Thanks to the weight vectors, we can calculate the cosine similarity^20^ and use this value to know if a word is specific to a cluster or if, on the contrary, it is shared by all.

In the average time series, we show in cool colors (dark blue, light blue) those segments of the time series that are cluster-specific, and in warm colors (red and orange), those segments that are similar across all clusters. Basically, the concept is that words with a very high frequency of occurrence in one cluster and a very low frequency in the others appear in blue. If the word appears repeatedly in the cluster and in the other clusters, they are shown in orange or red. If the occurrence in the other clusters is medium, the color is green or yellow.

## 3. Results

### 3.1. Participant characteristics

Twenty-five subjects participated in this study, of whom fourteen were women, and eleven were men. Table 1 shows the characteristics of the participants identified by a random ID and including gender (M=Male; F=Female), age, BMI, HbA1c, diabetes treatment (MDI: Multiples doses of insulin; CSII. Continuous subcutaneous insulin infusion) and years of evolution of T1DM.

**Table 1.** Characteristics of the participants: ID, Gender (M=Male; F=Female), Age, BMI, HbA1c, Treatment (MDI: Multiples doses of insulin; CSII: Continuous subcutaneous insulin infusion), Years of evolution of T1DM.

| ID | Gender | Age | BMI | HbA1c | Treatment | Years T1DM |
| --- | --- | --- | --- | --- | --- | --- |
| HUPA001 | F | Q4 | Q2 | Q4 | CSII | Q3 |
| HUPA002 | M | Q4 | Q2 | Q2 | CSII | Q4 |
| HUPA003 | F | Q3 | Q1 | Q2 | CSII | Q2 |
| HUPA004 | M | Q2 | Q4 | Q3 | CSII | Q1 |
| HUPA005 | F | Q1 | Q2 | Q1 | CSII | Q4 |
| HUPA006 | M | Q1 | Q3 | Q3 | CSII | Q2 |
| HUPA007 | M | Q2 | Q4 | Q1 | CSII | Q1 |
| HUPA008 | F | Q1 | Q4 | Q4 | CSII | Q1 |
| HUPA009 | F | Q3 | Q2 | Q3 | CSII | Q4 |
| HUPA010 | F | Q3 | Q1 | Q1 | CSII | Q2 |
| HUPA011 | F | Q2 | Q2 | Q3 | CSII | Q4 |
| HUPA014 | F | Q4 | Q3 | Q4 | MDI | Q2 |
| HUPA015 | F | Q3 | Q1 | Q1 | MDI | Q1 |
| HUPA016 | F | Q2 | Q3 | Q1 | CSII | Q3 |
| HUPA017 | F | Q1 | Q1 | Q4 | MDI | Q3 |
| HUPA018 | F | Q2 | Q1 | Q2 | CSII | Q4 |
| HUPA019 | M | Q1 | Q3 | Q2 | MDI | Q1 |
| HUPA020 | M | Q3 | Q3 | Q4 | MDI | Q2 |
| HUPA021 | F | Q4 | Q3 | Q3 | MDI | Q1 |
| HUPA022 | M | Q4 | Q2 | Q1 | CSII | Q2 |
| HUPA023 | M | Q1 | Q1 | Q3 | MDI | Q1 |
| HUPA024 | M | Q3 | Q4 | Q4 | MDI | Q4 |
| HUPA025 | M | Q2 | Q4 | Q1 | CSII | Q3 |
| HUPA026 | F | Q4 | Q4 | Q2 | MDI | Q3 |
| HUPA027 | M | Q1 | Q1 | Q2 | MDI | Q3 |
| **Average** | F:14/25 | 38.3 | 24.4 | 7.4 | CSII:15/25 | 18.1 |

The median and mean age are 41.2 and 38.3 years respectively, with an age range of 18-60.8 years while 1^st^ and 3rd quartiles are 26.4 and 47.9 years. The median/ mean duration of diabetes are 15.2/ 18.1 years with a range of 0.8-39.5 years and 1^st^/ 3^rd^ quartiles are11.2/ 24.2 years. HbA1c median/ mean are 7.3/ 7.4% (range 6-9.7%) and 1^st^/ 3^rd^ quartiles are 7/ 7.8%. Body Mass Index (BMI) median/ mean are 24.2/ 24.4 (range 18.5-32.2) and 1^st^/ 3^rd^ quartiles are 22.3/ 26.3. Fifteen patients were on continuous insulin pump therapy and ten on multiple insulin doses per day. The CGM shows a mean blood glucose of 155 mg/dl, with a high glycemic variability (CV 36) and a time in hypo and hyperglycemia above target.

Table 2 presents the values of the sleep monitoring variables for the participants of the study. Although most of the participants have a sleep efficiency higher than 90% there are also 3 cases with a value close to 45% and other two cases with low efficiency values (58% and 68%). Sleep data from some participants, such as HUPA007 or HUPA008 was discarded, due to inconsistency in the reported data. Although most participants have a sleep efficiency higher than 90%, there are also 3 cases with a value close to 45% and the other two cases with low-efficiency values (58% and 68%). Sleep data from some participants, such as HUPA007 or HUPA008 was discarded, due to inconsistency in the reported data.

**Table 2:**
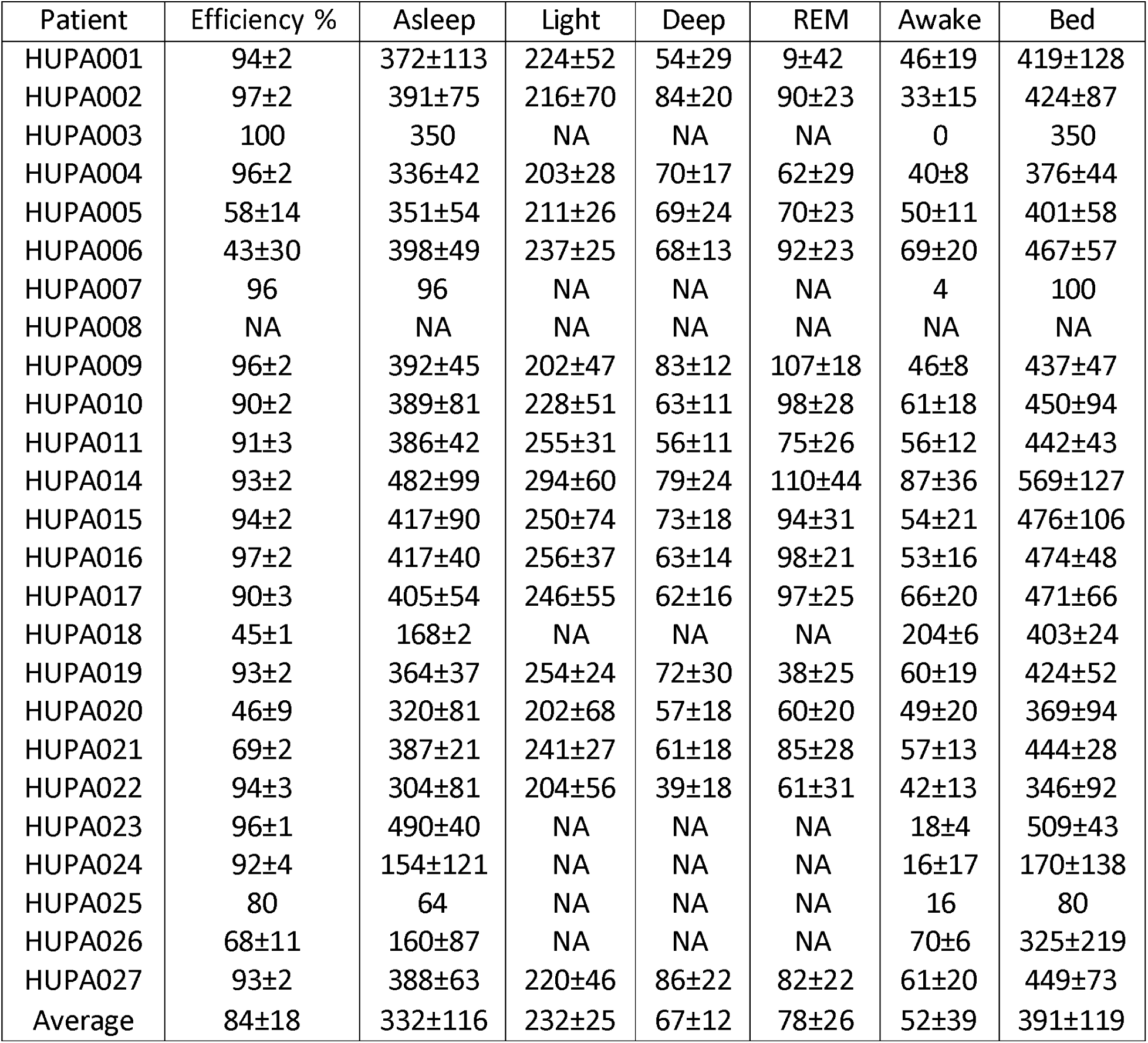
Average duration of sleep states with standard deviation (in minutes).

| Patient | Efficiency % | Asleep | Light | Deep | REM | Awake | Bed |
| --- | --- | --- | --- | --- | --- | --- | --- |
| HUPA001 | 94±2 | 372±113 | 224±52 | 54±29 | 9±42 | 46±19 | 419±128 |
| HUPA002 | 97±2 | 391±75 | 216±70 | 84±20 | 90±23 | 33±15 | 424±87 |
| HUPA003 | 100 | 350 | NA | NA | NA | 0 | 350 |
| HUPA004 | 96±2 | 336±42 | 203±28 | 70±17 | 62±29 | 40±8 | 376±44 |
| HUPA005 | 58±14 | 351±54 | 211±26 | 69±24 | 70±23 | 50±11 | 401±58 |
| HUPA006 | 43±30 | 398±49 | 237±25 | 68±13 | 92±23 | 69±20 | 467±57 |
| HUPA007 | 96 | 96 | NA | NA | NA | 4 | 100 |
| HUPA008 | NA | NA | NA | NA | NA | NA | NA |
| HUPA009 | 96±2 | 392±45 | 202±47 | 83±12 | 107±18 | 46±8 | 437±47 |
| HUPA010 | 90±2 | 389±81 | 228±51 | 63±11 | 98±28 | 61±18 | 450±94 |
| HUPA011 | 91±3 | 386±42 | 255±31 | 56±11 | 75±26 | 56±12 | 442±43 |
| HUPA014 | 93±2 | 482±99 | 294±60 | 79±24 | 110±44 | 87±36 | 569±127 |
| HUPA015 | 94±2 | 417±90 | 250±74 | 73±18 | 94±31 | 54±21 | 476±106 |
| HUPA016 | 97±2 | 417±40 | 256±37 | 63±14 | 98±21 | 53±16 | 474±48 |
| HUPA017 | 90±3 | 405±54 | 246±55 | 62±16 | 97±25 | 66±20 | 471±66 |
| HUPA018 | 45±1 | 168±2 | NA | NA | NA | 204±6 | 403±24 |
| HUPA019 | 93±2 | 364±37 | 254±24 | 72±30 | 38±25 | 60±19 | 424±52 |
| HUPA020 | 46±9 | 320±81 | 202±68 | 57±18 | 60±20 | 49±20 | 369±94 |
| HUPA021 | 69±2 | 387±21 | 241±27 | 61±18 | 85±28 | 57±13 | 444±28 |
| HUPA022 | 94±3 | 304±81 | 204±56 | 39±18 | 61±31 | 42±13 | 346±92 |
| HUPA023 | 96±1 | 490±40 | NA | NA | NA | 18±4 | 509±43 |
| HUPA024 | 92±4 | 154±121 | NA | NA | NA | 16±17 | 170±138 |
| HUPA025 | 80 | 64 | NA | NA | NA | 16 | 80 |
| HUPA026 | 68±11 | 160±87 | NA | NA | NA | 70±6 | 325±219 |
| HUPA027 | 93±2 | 388±63 | 220±46 | 86±22 | 82±22 | 61±20 | 449±73 |
| Average | 84±18 | 332±116 | 232±25 | 67±12 | 78±26 | 52±39 | 391±119 |

### 3.2. Sleep and glycemic control characteristics

Based on the National Sleep Foundation’s consensus recommendations for sleep quality assessment,^21^ we are going to employ three different criteria for defining sleep quality:

- Sleep efficiency: the ratio of total sleep time to time in bed.^10^
- WASO or Wake After Sleep Onset.
- The number of awakenings during night.

Table 3 shows the sleep characteristics of the participants. Sleep quality was categorized as poor if at least two of three criteria were met, i.e., sleep efficiency *<* 85% or Wake After Sleep Onset (WASO) *>* 40 min or number of awakenings *>* 4.

**Table 3.** Participants Sleep quality characterization categorized as ‘poor’ if at least two of three criteria were met, i.e., sleep efficiency *<* 85% or WASO *>* 40 min or number of awakenings *>* 4, based on the National Sleep Foundation’s consensus recommendations for sleep quality assessment.

| Participant | Total | Sleep Quality |  | Sleep Time | Sleep Efficiency | WASO | Awakenings |
| --- | --- | --- | --- | --- | --- | --- | --- |
| *ID* | *Nights* | *Good* | *Poor* | *Avg (hours)* | *Avg (%)* | *Avg (min)* | *Avg per night* |
| HUPA001P | 13 | 3 | 10 | 6.81±2.11 | 94.08±2.14 | 42.38±13.93 | 5±2.24 |
| HUPA002P | 11 | 8 | 3 | 7.07±1.45 | 97.45±1.63 | 33.36±14.53 | 2.73±1.9 |
| HUPA003P | 12 | 7 | 5 | 6.35±1.99 | 93.08±2.81 | 45.25±22.8 | 3.25±1.76 |
| HUPA004P | 10 | 6 | 4 | 6.22±0.75 | 95.8±1.81 | 40.1±7.75 | 3.2±1.4 |
| HUPA005P | 8 | 0 | 8 | 6.69±0.97 | 58.5±14.41 | 50.38±10.68 | 3.5±1.51 |
| HUPA006P | 6 | 0 | 6 | 7.54±0.98 | 47.67±33.68 | 68.33±22.44 | 6.33±3.33 |
| HUPA007P | 13 | 8 | 5 | 6.06±0.68 | 94.08±2.1 | 37.92±7.76 | 2.69±1.49 |
| HUPA011P | 13 | 1 | 12 | 7.37±0.72 | 90.92±3.43 | 55.92±11.84 | 4.23±1.83 |
| HUPA014P | 12 | 1 | 11 | 9.48±2.12 | 92.67±2.39 | 86.75±35.94 | 5.08±2.5 |
| HUPA015P | 13 | 3 | 10 | 7.94±1.76 | 93.69±1.55 | 54.23±21.19 | 3.69±1.49 |
| HUPA016P | 13 | 2 | 11 | 7.91±0.8 | 96.54±2.22 | 53.08±16.17 | 0.15±0.38 |
| HUPA017P | 13 | 1 | 12 | 7.85±1.11 | 90.31±3.35 | 66±19.94 | 4.85±1.68 |
| HUPA018P | 11 | 0 | 11 | 7.67±0.69 | 46.91±5.49 | 51.64±9.64 | 4.09±1.14 |
| HUPA019P | 6 | 0 | 6 | 7.06±0.87 | 93±1.67 | 60.33±19.25 | 6.17±5 |
| HUPA020P | 10 | 0 | 10 | 6.14±1.57 | 46.5±9.16 | 48.7±19.97 | 3.7±1.25 |
| HUPA021P | 8 | 0 | 8 | 7.4±0.47 | 69.38±2.2 | 56.88±13.29 | 4.62±1.85 |
| HUPA022P | 14 | 8 | 6 | 5.76±1.54 | 93.93±2.97 | 41.86±13.4 | 3.5±1.56 |
| HUPA023P | 10 | 0 | 10 | 7.93±1.33 | 95.5±1.18 | 55.1±9.35 | 4.8±1.4 |
| HUPA024P | 6 | 1 | 5 | 5.9±1.38 | 92±4.47 | 56.17±16.34 | 1.17±2.86 |
| HUPA025P | 12 | 1 | 11 | 7.32±1.01 | 92.08±3 | 55.75±13.14 | 4.17±2.52 |
| HUPA026P | 16 | 0 | 16 | 8.22±0.71 | 60.75±5.86 | 55.88±12.1 | 5.31±1.82 |
| HUPA027P | 13 | 4 | 9 | 6.52±0.81 | 93.38±2.33 | 45.85±11.12 | 3.23±1.54 |
| **Overall** | **243** | **54** | **189** | **7.15±1.17** | **83.1±4.99** | **52.81±15.57** | **3.88±1.93** |

Table 4 shows the overnight glycemic characteristics of the participants. The sleep characteristics of the participants show large inter-individual differences, and only 48% of the patients have a good overall sleep quality, although the mean sleep duration is not low (mean of 7.15 hours).

**Table 4.** Overnight glycemic characteristics in individual participants

| user_id | Nights | Mean_glucose | SD | CV | TR | T Hyper | T Hypo |
| --- | --- | --- | --- | --- | --- | --- | --- |
| HUPA001P | 13 | 181.71±32.27 | 67.12±10.97 | 37.27±5.07 | 54.21±13.77 | 43.54±14.74 | 2.25±3.13 |
| HUPA002P | 11 | 113.68±30.59 | 50.18±15.85 | 44.72±11.52 | 60.94±16.59 | 15±17.3 | 24.07±18.09 |
| HUPA003P | 12 | 139.88±22.85 | 55.38±16.57 | 38.98±5.56 | 69.66±12.53 | 22.69±14.68 | 7.65±7.24 |
| HUPA004P | 10 | 178.75±44.75 | 74.15±23.86 | 44.07±14.86 | 44.37±19.36 | 43.7±22.39 | 11.93±13.73 |
| HUPA005P | 8 | 151.06±23.74 | 43.05±14.68 | 29.29±11.12 | 69.09±16.96 | 27.21±18.19 | 3.7±4.45 |
| HUPA006P | 6 | 212.05±97.73 | 62.41±36.48 | 35.97±20.23 | 46.76±28.69 | 48.68±29.74 | 4.56±5.55 |
| HUPA007P | 13 | 173.64±29.53 | 73.14±14.31 | 42.53±8.47 | 46.28±17.35 | 45.07±17.61 | 8.65±8.49 |
| HUPA011P | 13 | 159.3±19.37 | 54±8.38 | 34.09±4.97 | 65.47±10.23 | 31.96±11.56 | 2.57±3.52 |
| HUPA014P | 12 | 186.47±19.12 | 68.55±22.54 | 36.68±11 | 44.96±10.21 | 50.83±11.34 | 4.2±3.77 |
| HUPA015P | 13 | 165.9±20.72 | 65.03±10.67 | 39.33±5.08 | 57.68±12.01 | 38.6±13.38 | 3.72±3.22 |
| HUPA016P | 13 | 157.32±45.81 | 67.24±20.44 | 43.51±12.62 | 51.16±18.54 | 36.1±22.75 | 12.74±10.33 |
| HUPA017P | 13 | 198.39±27.38 | 62.06±16.51 | 31.67±8.14 | 37.39±16.94 | 60.25±18.3 | 2.35±3.36 |
| HUPA018P | 11 | 144.12±35.97 | 62.64±17.93 | 43.6±5.62 | 49.73±11.83 | 31.77±18.56 | 18.5±14.91 |
| HUPA019P | 6 | 159.97±17.59 | 54.82±5.05 | 34.4±2.64 | 59.14±11.14 | 36.85±11.92 | 4.01±2.24 |
| HUPA020P | 10 | 193.99±29.73 | 72.11±18.31 | 37.23±7.74 | 44.92±14.04 | 51.29±15.31 | 3.78±3.88 |
| HUPA021P | 8 | 141.27±14.84 | 44.83±5.46 | 31.92±4.32 | 74.07±9.15 | 22.78±10.68 | 3.14±5.71 |
| HUPA022P | 14 | 111.12±23.51 | 31.58±7.55 | 29±6.96 | 79.57±11.25 | 5.01±8.62 | 15.42±13.42 |
| HUPA023P | 10 | 132.89±21.29 | 38.98±7.22 | 29.53±4.81 | 78.92±12.69 | 18.05±14.39 | 3.03±4.47 |
| HUPA024P | 6 | 157.51±31.58 | 56.53±18.8 | 37.25±14.74 | 57.63±9.82 | 35.17±15.25 | 7.19±10.08 |
| HUPA025P | 12 | 113.51±14.56 | 36.46±9.21 | 32.09±7.44 | 79.58±11.24 | 7.42±6.85 | 12.99±8.94 |
| HUPA026P | 16 | 133.64±24.6 | 57.68±20.78 | 42.86±11.98 | 60.7±21.91 | 22.25±17.37 | 17.05±13.76 |
| HUPA027P | 13 | 121.24±21.16 | 30.73±8.24 | 25.82±7.92 | 87.04±18.09 | 8.36±18.09 | 4.6±4.91 |
| Overall | 243 | 155.79±29.49 | 55.85±14.99 | 36.45±8.76 | 59.97±14.74 | 31.94±15.86 | 8.1±7.6 |

Of the 243 nights analyzed, 77% (*n* = 189) were of poor quality and 33% (*n* = 54) of good quality. It should be noted that eight patients had no night with good sleep quality. The factor that most determined poor sleep quality was the duration of nighttime awakenings, with a mean of 52.81 minutes.

### 3.3. Association between sleep quality and blood glucose

In Figure 2, we can see the upper triangular of the correlation matrix for different variables recorded in this study displayed as a correlogram, with colors red/ blue for showing negative/ positive correlation and low/ high intensity indicating the absolute value of the correlation. In addition to the expected correlations, we can point out the following facts:

- Both poor and good sleep qualities have no significant relationship with any other variable.
- A positive correlation of 0.8 was found between the standard deviation of sleep efficiency and the standard deviation of mean blood glucose.
- In addition, the standard deviation of sleep efficiency has a positive correlation (0.66/ 0.62/ 0.65/ 0.61) with the standard deviation of glucose, the coefficient of variation, the time in range, and the time in hyperglycemia.
- There is a positive correlation (0.55) between the coefficient of variation and the mean time spent in hypoglycemia.

**Figure 2:**
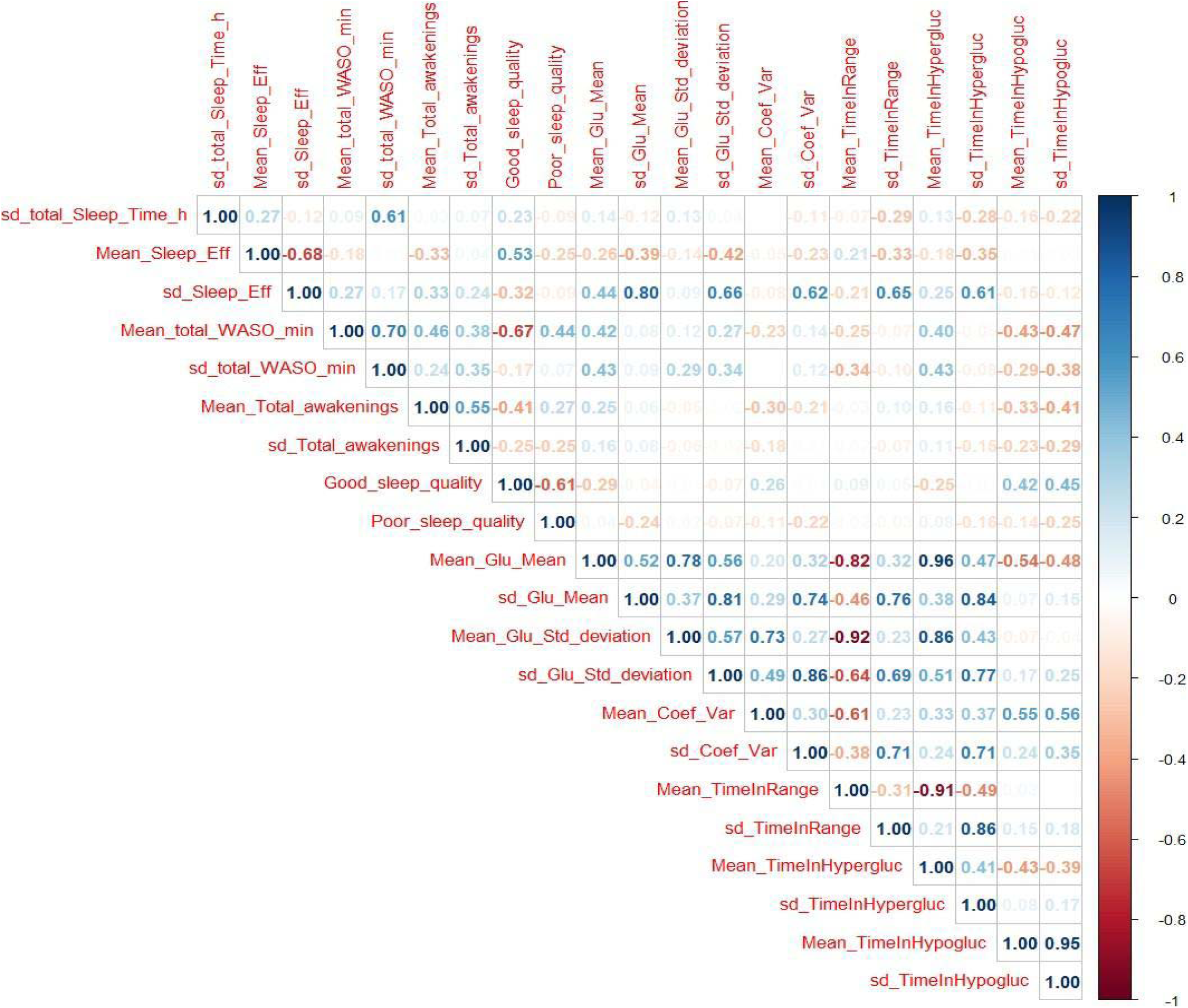
Correlation matrix for all the variables recorded in this study

As we have seen, from Figure 2 we cannot draw clearly significant relationships among sleep quality, efficiency and glucose variables, but what about sleep patterns?

We have used the clustering techniques described in section 2.2 to find patterns in sleep behavior. We have experimented with a different number of clusters, having found *k* = 4 to be the best option, showing a clear relationship between sleep structure and glucose variables. With *k* = 3, we have a cluster with very broad glucose patterns, whereas with *k* = 5, the relationship between sleep structure and the different glucose variables is confirmed with no additional information, remaining the main clusters the same.

In the left column of Figure 3, we can see the four sleep clusters, while the right column shows the average glucose behavior in the days corresponding to each cluster. Recall that in the second case, the specificity of the glucose patterns is shown with intense blue color, while the patterns common to all clusters are shown in red. Because we take glucose level samples every 15 minutes, we have 96 samples per day, and in the horizontal axis we can see marked the main hourly correspondences. To correctly compare the sleep clusters, we show a total of 44 possible states per night (horizontal axis, on the left column).

**Figure 3:**
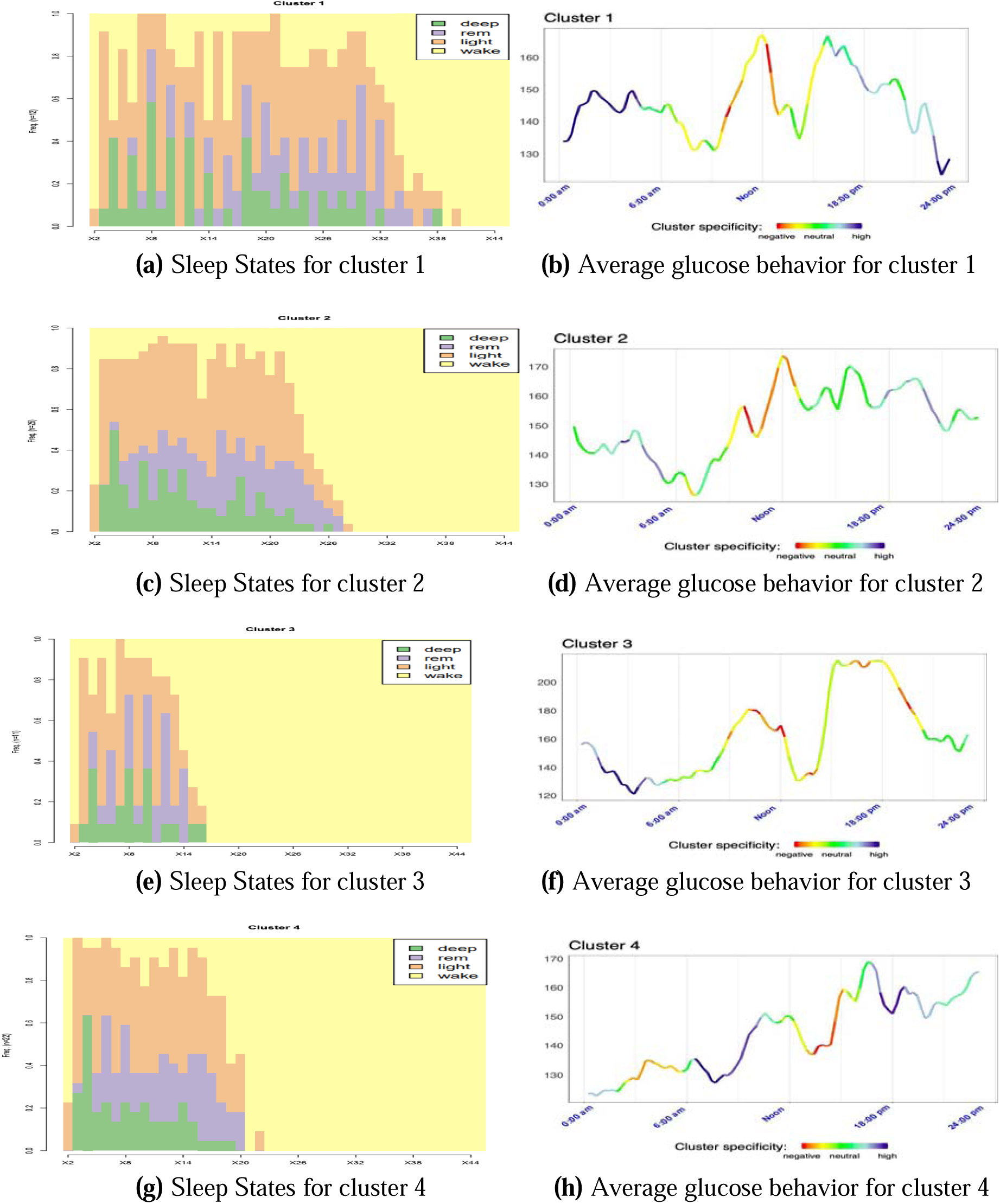
Structure of the sleep states and average glucose for the different clusters with k=4.

We have also calculated the Shannon entropy^22^ for the 4 clusters resulting in the following order (from highest entropy to lowest): 1,3, 2, and 4. Shannon’s entropy can give us an idea of how ordered or disordered the sleep has been. Higher entropy means higher disorder. Moreover, Figure 4 shows the results for each cluster of the main glucose variables. So, from the parallel observation of both Figures (3 and 4) several conclusions arise:

- Clusters 1 and 2 (figures 3a and 3c) have the longest sleep state sequences, the highest nocturnal glucose levels, and a maximum peak around noon. Clusters 3 and 4 (figures 3e and 3g), with the shortest sleep state sequences have the lowest nocturnal glucose levels and a maximum peak around sunset. Besides, these two clusters are also the two clusters with the lowest time in hypoglycemia (figure 4b). In the case of cluster 3, this is mainly because it is the cluster with the longest time in hyperglycemia (figure 4c), while cluster 4 is the cluster with the longest time in range (figure 4a).
- Cluster 1, with the highest Shannon entropy and the longest sequence of states (40), has as a specific characteristic a very pronounced drop in glucose levels prior to the night and at the same time the most accentuated nocturnal rise (displayed in dark blue in figure 3b). In addition, it is the cluster with the lowest time in range (figure 4a), the highest time in hypoglycemia (figure 4b), coefficient of variation (figure 4d) and standard deviation (figure 4f).
- Cluster 3, with the shortest sleep sequence (16 states), has the unique characteristic of a pronounced drop in glucose level during the night (figure 3f). It is also the cluster with the highest time in Hyperglycemia (figure 4c) and, therefore, the highest mean level of glucose (figure 4e).
- In Figure 4f, we can see that the lowest standard deviation corresponds to clusters 2 and 4. This could be related with the fact that these two clusters have the sleep state sequences with the lowest Shannon entropy.

**Figure 4:**
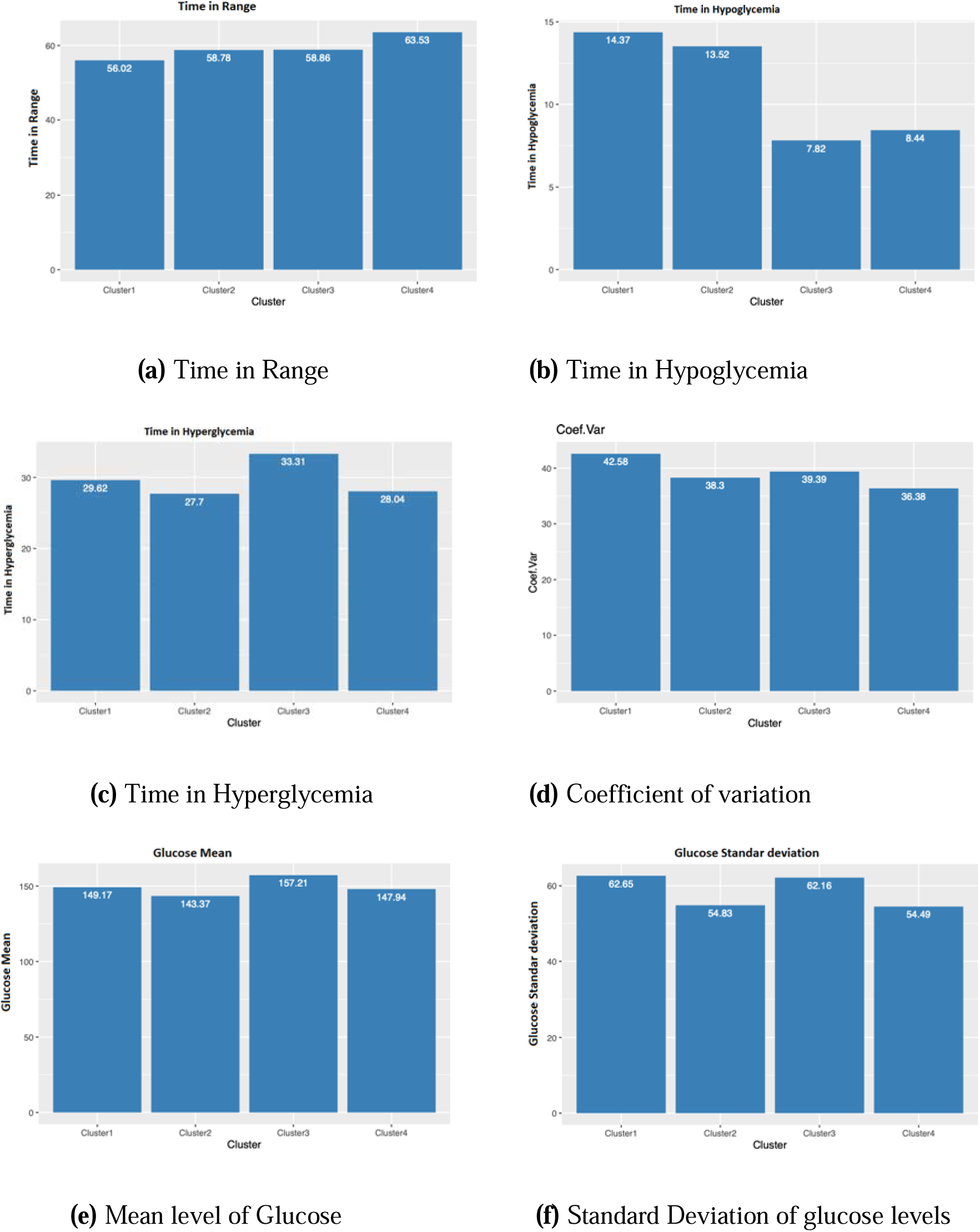
Characteristics of the different clusters. Results for 4 clusters

The shortest time in hyperglycemia and therefore lower mean glucose level (figures 4c and 4e) corresponds to cluster 2, which has a low level of Shannon entropy and a medium length of the sleep state sequence (figure 3c).

## 4. Discussion

This study shows that in a randomly selected population of adults with type 1 diabetes, poor sleep quality as measured by sleep actigraphy is very frequent as it occurs in 77% of the nights analyzed (189/243). In a previous epidemiological study using the Pittsburgh survey to measure subjective sleep quality^1^ in a sample of 222 patients, they found that 41% have poor sleep quality (Pittsburgh index >5). According to the experimental data of our study, variability in sleep quality in adults with type 1 diabetes is associated with more significant variability in nocturnal blood glucose levels.

To our best knowledge, this is the first study using artificial intelligence statistical techniques to analyze the relationship between sleep structure and times in normo-, hypo-, and hyperglycemia and to show that better sleep structure is associated with longer time in glycemic range during that day.

These findings confirm the results of a previous study in 20 adult patients^23^ showing that poor sleep quality is associated with greater glycemic variability. However, they found no association between sleep quality and time in range. This study only analyzes the relationship between sleep quality and glycemia with a linear mixed-effects model.

Other previous studies that do not use continuous glucose monitoring also suggest that sleep disturbances worsen glucose control:

- Patients with short sleep duration (<6.5 hours) have higher HbA1c than patients with longer sleep duration (>6.5 hours).^24^
- Increased time in deep sleep phase has been correlated with lower HBA1c and less time in nocturnal hypoglycemia.^25^
- Social jet lag (major changes in the duration and timing of sleep between weekdays and holidays) has been associated with worse chronic metabolic control.^26^

Some studies demonstrate the influence of sleep quality or duration on glycemic control in children. However, the findings among the different authors are not the same: it has been reported that a longer duration of the light sleep phase is associated with higher mean daily blood glucose, more episodes of hyperglycemia, and higher HbA1c,^4^ and that increased nocturnal awakenings^27^ correlate with high glycemic variability.

Most of these studies have limitations in that they have been conducted on a small number of patients, some have only used subjective sleep assessments, and only two studies in adults have simultaneously performed continuous glucose monitoring and polysomnography. There are several possible mechanisms involved in poorer glycemic control:^28^

- Decreasing the duration of the REM phase would produce lower nocturnal glucose consumption, given that in the cerebral REM phase, glucose consumption is similar to awake. In contrast, in the cerebral non-REM phase, glucose consumption is much lower.
- In the general population and in patients with diabetes, sleep deprivation and fragmentation and decreased deep sleep are associated with decreased insulin sensitivity, possibly mediated by increased cortisol and Growth hormone (GH) levels. In patients with T1DM, higher nocturnal levels of growth hormone, adrenaline, ACTH, and cortisol have been reported than in the control population.^3^
- In an experimental study, partial restriction of a single night of sleep (4 hours) decreased peripheral insulin sensitivity measured by the hyper insulinemic euglycemic clamp in patients with DM1.^29^

Although it has been proposed that continuous glucose monitoring may alter sleep quality due to hyper or hypoglycemia alarms, in this study, the CGM did not have alarms, so the likelihood of interference of the CGM on sleep quality is very low.

Finally, sleep disturbances could worsen glycemic control by an indirect mechanism related to patients’ behavior and cognitive functions. An association has been described in children and teenagers between shorter duration of sleep and a decrease in the frequency of self-monitoring and insulin bolus administration.^30^

## 5. Conclusion

To our best knowledge, our work is the first study that by means of artificial and intelligence statistical techniques has found a relationship between sleep structure and times in normo-, hypo-, and hyperglycemia. Our main conclusion is that better sleep structure is associated with longer time in glycemic range.

Future studies are needed to confirm these findings in a larger patient population along with investigation of the mechanisms involved in the decreased time in range and increased glycaemic variability caused by poor sleep quality. We believe that sleep disturbances should be a factor to be assessed in the clinical practice of patients with type 1 diabetes and that strategies should be designed to treat these disturbances.

## Data Availability

All data produced in the present study are available upon reasonable request to the authors

## 6. Acknowledgements

This work has been supported by:

- Fundación Eugenio Rodriguez Pascual 2019 grant Desarrollo de sistemas adaptativos y bioinspirados para el control glucémico con infusores subcutáneos continuos de insulina y monitores continuos de glucosa (Development of adaptive and bioinspired systems for glycemic control with continuous subcutaneous insulin infusors and continuous glucose monitors).
- The Spanish Ministerio de Innovación Ciencia y Universidad grant RTI2018095180-B-I00.
- Madrid Regional Goverment FEDER grants B2017/BMD3773 (GenObIA-CM) and Y2018/NMT-4668 (Micro-StressMAP-CM).

## 7. Author Disclosure Statemen

The authors declare no conflict of interest.

